# Association of Suzetrigine With Postoperative Outcomes Versus Opioid Analgesics: Propensity-Matched Study

**DOI:** 10.64898/2026.03.29.26349666

**Authors:** Abhinav S. Verma, Vasudha Sharma, Rishi Chowdhary, Anjali Pathak, Suha Soni, Vishalika Gandhari, Terence Hillery, Rishab Gupta

## Abstract

**Background:** Perioperative pain management modality potentially influences psychiatric morbidity and healthcare utilization. The opioids have been most commonly used for managing postoperative pain and carry a high degree of risk for creating mood disorders, anxiety, sleep disturbances, and healthcare burdens. A novel non-opioid analgesic, Suzetrigine, may be able to effectively manage postoperative pain without some of the psychological and economic risks that come from the use of opioids. In this study, we measured psychiatric outcomes and emergency department (ED) usage among postoperative patients who received either suzetrigine or opioids.

**Methods:** This was a retrospective cohort study using the TriNetX US Collaborative Network, encompassing 64 healthcare organizations. Adult patients (> Age 18 years) who underwent surgery and received suzetrigine were compared with patients who underwent surgery and received opioids. Propensity score matching (1:1) performed to match cohorts based on demographic factors (age, gender, racial/ethnic status), social determinants of health (ICD-10 Z55-Z65), family histories of substance abuse and psychiatric disorders (Z81.x), surrogate measures of prior healthcare utilization, and pre-existing clinical severity using Elixhauser-Charlson comorbidity proxies (hypertensive diseases [I10-I15], diabetes mellitus [E08-E13], ischemic heart disease [I22-I25], and chronic pulmonary disease [J42-J47]). Matching also included behavioral risk factors (tobacco use and physical inactivity) and body mass index (BMI). Following matching, there were 2,221 patients in each cohort. The primary outcome assessed within one year after surgery was ED utilization, depression, anxiety, post-traumatic stress disorder (PTSD) and sleep disorders. Risk estimates and survival analyses were used to compare the outcomes.

**Results:** In propensity-matched analyses, suzetrigine use was associated with a reduction in multiple psychiatric outcomes and healthcare utilization compared to opioid analgesics. There was less ED utilization in the suzetrigine cohort (5.9% v 13.1%, RR 0.45, p< .001). The psychiatric outcomes were also lower in the suzetrigine cohort than the opioid cohort, including depression (3.1% v 4.7%, RR 0.65, p= .005), anxiety (4.7% v 7.2%, RR 0.65, p< .001), PTSD (0.5% v 1.4%, RR 0.36, p= .002), and sleep disorders (4.2% v 6.0%, RR 0.71, p= .008). The survival analysis suggested an earlier onset of psychiatric diagnosis among the opioid recipients.

**Conclusion:** In a matched real-world cohort of surgical patients, suzetrigine use was associated with lower short-term rates of selected postoperative outcomes compared with opioid analgesics.

## INTRODUCTION

Postoperative pain is one of the major factors influencing a patient’s recovery from a surgical procedure and is a known primary reason for unplanned care, delayed mobilization, excessive postoperative opioid consumption, and patient-reported dissatisfaction with their recovery process. ^1^. Opioids have been the first-line treatment for perioperative pain for decades, but increasing awareness among both physicians and patients about their long-term side effects drives the search for better alternatives. ^2^

Several reports have emphasized the need to prevent the long-term risks associated with postoperative opioid exposure. In a large prospective cohort study of patients discharged on opioids, the total morphine-equivalent dose of opioids that was prescribed at discharge was approximately two times greater than the amount consumed during the first week following discharge, indicating excessive circulating opioid levels and the need for opioid stewardship. ^3^ Consistent with concerns regarding the long-term risks associated with postoperative opioid exposure, large U.S. claims data have shown that postoperative opioid prescription sizes and the number of individuals who develop new persistent opioid use have declined over time but continue to hold importance and are predictable in part due to prior baseline characteristics, including controlled substance use patterns. ^4^. Observational studies indicate that new persistent opioid use after surgery can increase healthcare expenditures by up to $17,000 per patient within the year following surgery, underscoring the broader economic burden associated with opioid-based postoperative pain management. ^5^

One of the latest and most promising options, Suzetrigine (VX-548), represents a novel mechanism of action as a non-opioid, peripherally-acting analgesic through selective inhibition of the voltage-gated sodium channel NaV1.8, which provides the potential to provide effective pain relief without activating the central opioid receptors. Both the underlying science and translational aspects of NaV1.8 inhibition and the safety assessments of suzetrigine from key studies ^6^ are reviewed. Clinical reviewers discuss suzetrigine’s post-approval positioning for moderate-to-severe acute pain. ^7^ Additionally, analyses suggest that although suzetrigine is more expensive than generic opioid analgesics, studies indicate that the medication may be cost-effective or even cost-saving over a patient’s lifetime if it reduces opioid-related complications such as opioid use disorder. (Rind et al., 2025; Nikitin et al., 2025)

Randomized phase II and phase III studies of postoperative pain demonstrated suzetrigine to be more efficacious compared to placebo over relatively short periods of time. Trials also compare the analgesic effect of suzetrigine to its opioid comparators, demonstrating the promising effects of suzetrigine over traditional opioids. Despite the thoroughness of these trials in comparing the pharmacologic efficacy of suzetrigine vs placebo and traditional opioids, several deficiencies remain. ^8^

The trials conducted so far did not measure long-term outcomes such as the incidence of depression/anxiety/PTSD, sleep disorders, or acute care utilization in the subsequent year.^9^ Observational perioperative opioid outcomes literature continues to be limited by uncontrolled confounding by indication and baseline risk factors, including socioeconomic/psychosocial status and burden of comorbid conditions, which systematic reviews and meta-analyses have identified as being associated with the development of persistent opioid use and related outcomes. ^10^

Despite evolving pain management strategies, substantial knowledge gaps remain regarding how systemic non-opioid analgesic classes perform relative to opioid-containing regimens in real-world practice, as well as which clinically meaningful downstream outcomes extend beyond the immediate assessment of acute pain intensity. Notably, psychiatric and sleep-related disturbances are increasingly recognized as important postoperative complications across diverse surgical populations. ^11,12^ Moreover, observational perioperative literature has demonstrated associations between prolonged opioid exposure and adverse mood and sleep outcomes in other clinical contexts ^13^ Thus, additional longitudinal comparative studies of postoperative analgesic strategies and downstream psychiatric/sleep/utilization outcomes are required with carefully measured confounding and clearly defined operationalized baseline covariates to better understand the relationships between postoperative analgesic strategy and psychiatric, sleep, and utilization outcomes in the longer term.

We assessed whether postoperative pain management with suzetrigine, compared with opioid analgesics, was associated with differences in subsequent diagnoses of depression, anxiety, PTSD, sleep disorders, and ED visits over 1 year in a large, multicenter EHR network after propensity score matching.

## METHODS

We relied upon the TriNetX U.S. Collaborative Network. The network comprises over sixty hospitals in the United States and incorporates de-identified EHR information from them. In addition to providing real-time access to data regarding encounters, diagnoses, procedures, medications, etc., it has analytic capabilities to support cohort development and comparison of outcomes. ^14^

### Study Design and Population

Suzetrigine (Journavx, formerly VX-548), a novel, non-opioid sodium channel NaV1.8 blocker for acute pain, received FDA approval as of January 30, 2025.^15^ Thus, in order to establish consistent time frames for drug exposure classification, our patient cohort included only those who received suzetrigine or opioid analgesics subsequent to January 30, 2025.

The sample for this retrospective cohort study consisted of adults (≥ 18 years old) undergoing surgery for whom either suzetrigine or opioid analgesics were administered postoperatively for pain management. The index event was the first qualifying surgery (Current Procedural Terminology 1003143) that met the exposure criteria. Exposure to suzetrigine was defined as administration of suzetrigine at any time between one day before and seven days after the qualifying surgery. Exposure to opioid analgesics was defined as administration of any opioid (e.g., codeine, hydrocodone, oxycodone, morphine, tramadol, or other opioids) at any time one day before and seven days after the surgery.

In order to limit confounding resulting from prior opioid exposures, all patients were excluded if they had an opioid prescription documented or were diagnosed with an opioid-related disorder (ICD-10 F11); other psychoactive substance use disorders (F10-F19); chronic pain not otherwise classified (G89.2); or documented exposure to both suzetrigine and an opioid during the exposure window (any time before January 30, 2025). This limitation was applied equally to both cohorts so that each cohort would have a similar level of risk for adverse opioid related outcomes. All remaining patients who were at risk for the outcomes at baseline were included in the study.

### Follow-up and Outcomes

Follow-up commenced one day after the index surgery and continued for 365 days. Three primary outcome measures were: (1) ED visits; (2) new diagnoses of depression, anxiety, PTSD, and sleep disorders. To eliminate immortal time bias, patients with documentation of a primary outcome measure occurring prior to the initiation of follow-up were eliminated from the respective primary outcome measure analysis. Definitions for the primary outcome measures were developed utilizing ICD-10 diagnosis codes and prescription records contained within the TriNetX database.

### Propensity Score Matching

To account for confounders, a PSM was performed using the TriNetX analytics platform. The PSM employed a logistic regression model to estimate propensity scores. The model included demographic covariates (age at index, sex, race/ethnicity); social determinants of health (ICD-10 Z55-Z65); lack of physical exercise (Z72.3); tobacco use (Z72.0); hypertensive diseases (I10-I15); diabetes mellitus (E08-E13); ischemic heart disease (I20-I25); family history of mental and behavioral disorders (Z81.x); and BMI. The PSM employed a greedy 1:1 nearest neighbor algorithm without replacement, with a 0.1 pooled-standard-deviation caliper (TriNetX default), to match patients who received suzetrigine to patients who received opioid analgesics. ^16^ The balance of covariates was evaluated using standardized mean differences (SMD), with values ≤0.10 indicating adequate balance. Following the completion of the PSM, 2221 patients remained in each cohort.

### Statistical Methods

Descriptive statistics for baseline characteristics were calculated using means and standard deviations for continuous variables, and frequency counts and percentages for categorical variables. SMDs were used to describe the difference between the matched cohorts. Risk ratios (RR) and 95% confidence intervals (CI) for each outcome were estimated using log-binomial regression, and p values < 0.05 were determined to be statistically significant. The RR, CI, and p value were calculated for each outcome by the TriNetX platform, which uses the matched cohorts to calculate the number of events, RR, and CI for each outcome.

## RESULTS

### Cohort Identification

Of the 64 healthcare systems within the TriNetX US Collaborative Network, 2,325 adult patients that underwent surgical procedures and were prescribed suzetrigine (cohort A) and 1,273,084 adult patients that were prescribed an opioid analgesic (cohort B) were initially identified. Following application of the inclusion/exclusion criteria and 1:1 propensity score matching of the two cohorts, there were 2,221 patients in each cohort. The standardized mean difference (SMD) for each of the covariates used in matching indicated that each SMD was less than 0.10, suggesting that the cohorts were well balanced.

### Baseline Characteristics

The demographic profiles of the two cohorts were similar. In terms of age, the average age of the patients in both cohorts was approximately 52 years. Approximately 54% of the patients in both cohorts were female. The proportion of patients in each cohort that reported being Hispanic/Latino was low. There were few significant differences in the prevalence of pre-existing medical conditions such as diabetes mellitus and hypertensive disease between the two cohorts. However, the prevalence of pre-existing mental health diagnosis was somewhat higher in cohort A than in cohort B (e.g., 17.0% vs. 11.0% for anxiety disorders). On average, the body mass index (BMI) of the patients in cohort A was approximately 28.6 kg/m^2 while the BMI of the patients in cohort B was approximately 29.5 kg/m^2.

### Primary Outcome

A total of 132 (5.9%) of the 2,221 patients in the suzetrigine cohort visited the emergency department (ED) after their surgery compared to 292 (13.1%) of the 2,221 patients in the opioid cohort. This represents a risk ratio of 0.45 (95% Confidence Interval (CI): 0.37 – 0.55; p < .001) for ED visits post-surgery for suzetrigine compared to opioid. The time-to-event analysis demonstrated that suzetrigine recipients were at reduced risk of visiting the ED post-surgery compared to those receiving opioids, which is represented by a hazard ratio of 0.41 (95% CI: 0.32 – 0.52).

### Secondary Outcomes

Compared to the patients that used opioids, the suzetrigine cohort had lower rates of depression (3.1% vs. 4.7%, Risk Ratio 0.65 [95% CI 0.48 – 0.88]; p = 0.005) and anxiety (4.7% vs. 7.2%, Risk Ratio 0.65 [95% CI 0.51 – 0.83]; p < 0.001). Additionally, post-traumatic stress disorder (PTSD) was diagnosed in 0.5% of the suzetrigine patients compared to 1.4% of the opioid patients (Risk Ratio 0.36 [95% CI 0.18 – 0.70]; p = 0.002). Finally, sleep disorders were diagnosed in 4.2% of cohort A and 6.0% of cohort B (Risk Ratio 0.71 [95% CI 0.55 – 0.91]; p = 0.008).

The Cox Proportional Hazards Models replicated the crude Risk Ratio Analyses. The HRs for Depression, Anxiety and Sleep Disorders were 1.79, 1.64 and 1.71, respectively, which indicated that there was an increased hazard for each of these psychiatric outcomes in the Opioid Cohort as time passed. There were no additional Independent Predictors identified.

## DISCUSSION

A large retrospective cohort study of 4,442 patients identified through the TriNetX US Collaborative Network found that suzetrigine was associated with a decreased risk of emergency department utilization when compared to opioid analgesics (RR 0.45, 95% CI 0.37–0.55), and the results were consistent regardless of whether the subjects were propensity score-matched to adjust for baseline differences in the covariates. Additional secondary analyses indicated lower risks of depression, anxiety, post-traumatic stress disorder, and sleep disorders among subjects exposed to suzetrigine, and higher rates of prescription for opioids to treat opioid-use disorder. Time-to-event analyses yielded results that were similar in terms of both the magnitude and direction of the associations.

Since suzetrigine was introduced into clinical practice recently, prescribing may have been affected by clinician preference for drug, institutional availability of drug, formulary adoption, and other patient level variables that are not reflected well within the electronic health record (EHR). Therefore, while we attempted to address confounding through propensity score matching, this study has limitations due to confounding by indication and the associations described should be viewed as hypothesis generating, but not causal.

The recent perioperative literature has placed increased emphasis on minimizing opioid exposure while providing sufficient analgesia. Randomized trials and meta-analyses examining opioid-sparing strategies have consistently shown a reduction in opioid consumption with no loss of analgesic efficacy; however, the long-term impact of these strategies on psychiatric and healthcare utilization outcomes is not as clearly defined. ^17,18^ Studies based on large electronic health record databases have also documented an association between postoperative opioid exposure and subsequent diagnoses of mental health conditions and increased healthcare utilization. ^19,20^

This study used a large, geographically diverse, multicenter electronic health record network representing current clinical practices throughout the United States. The analytical strategy used 1:1 propensity score matching to equate demographic, comorbidity, social determinant of health, and baseline clinical proxy covariates that could produce confounding. Utilization of real-world data increased the external validity of the study and captured the usual clinical practices of perioperative prescribing in the hospitals. Moreover, the federated TriNetX platform facilitates the reproducible estimates of effect.

There are several limitations that should be considered. First, the identification of diagnoses and exposures occurred via International Classification of Diseases (ICD) and medication coding and therefore there is potential for misclassification. Second, even though propensity score matching equalized measured covariates, it is still possible to have residual confounding due to unmeasured covariates such as pain severity, intraoperative anesthesia regimen, unmeasured socioeconomic factors not represented by Z codes, and/or poor medication adherence. Third, detailed clinical information regarding the dosing and duration of the administered analgesics and postoperative pain scores were not available. Fourth, encounters with the healthcare system occurring outside the boundaries of the participating systems may not have been fully represented. Fifth, participating institutions within the federated EHR networks may disproportionately represent larger academic or integrated health systems, potentially limiting generalizability. Sixth, the observational design of this study does not allow for causal inference.

## CONCLUSIONS

In this large multicenter retrospective cohort study, exposure to suzetrigine during the perioperative period was associated with lower risks of emergency department utilization and several psychiatric diagnoses when compared to opioid analgesics after adjusting for baseline differences in the covariates via propensity score matching. Further prospective studies including randomized controlled trials when feasible will be necessary to further evaluate these associations.

## Supporting information

Strobe Checklist

## Data Availability

Data used in this study were obtained from the TriNetX Research Network. Due to data use agreements and the de-identified nature of the dataset, individual-level data cannot be shared. Aggregated data supporting the findings are available from the corresponding author upon reasonable request, subject to TriNetX policies.

https://live.trinetx.com/

## FIGURE AND TABLE LEGEND

**Figure 1.**
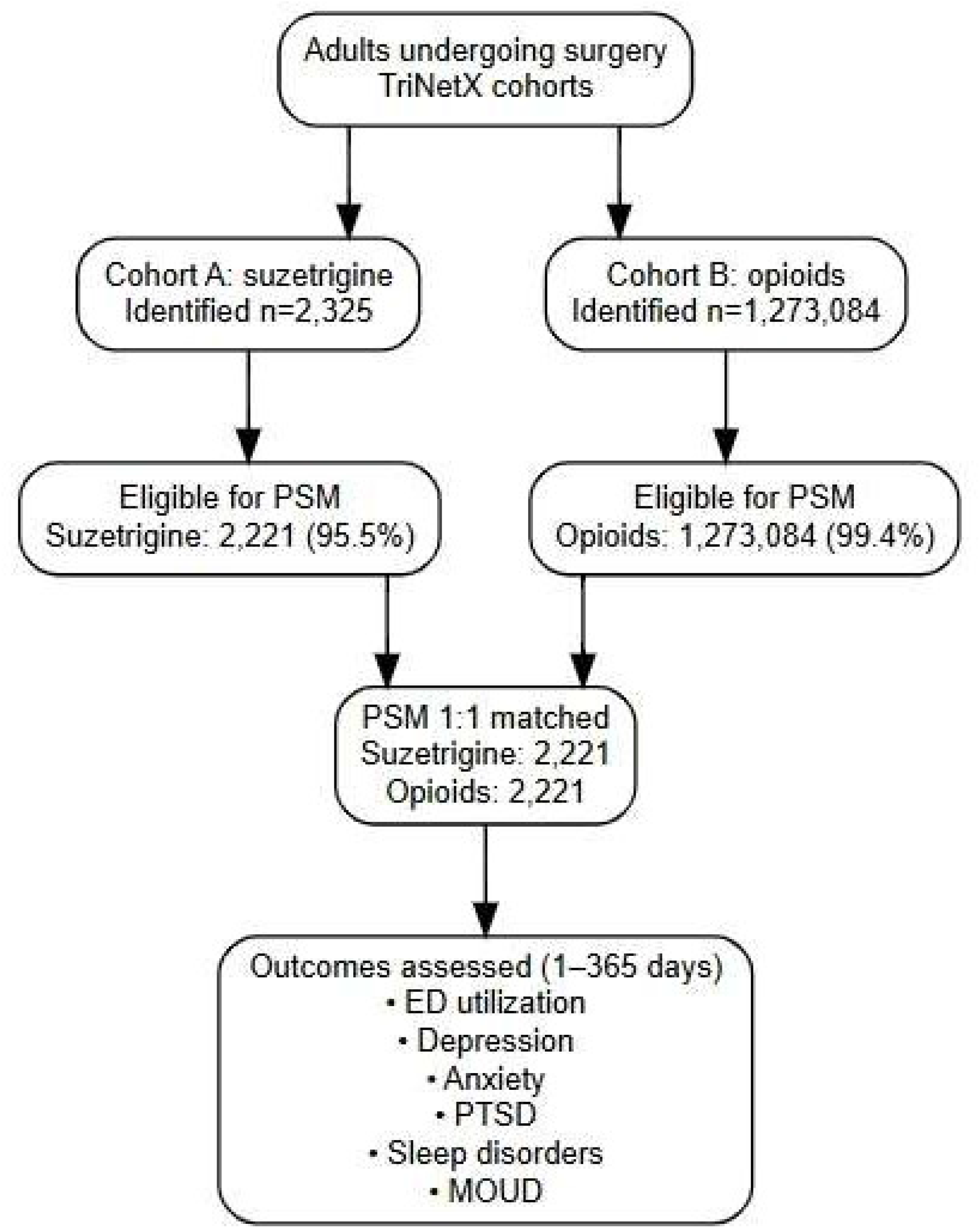
CONSORT diagram of study/control cohorts’ selection flow.

**Figure 2.**
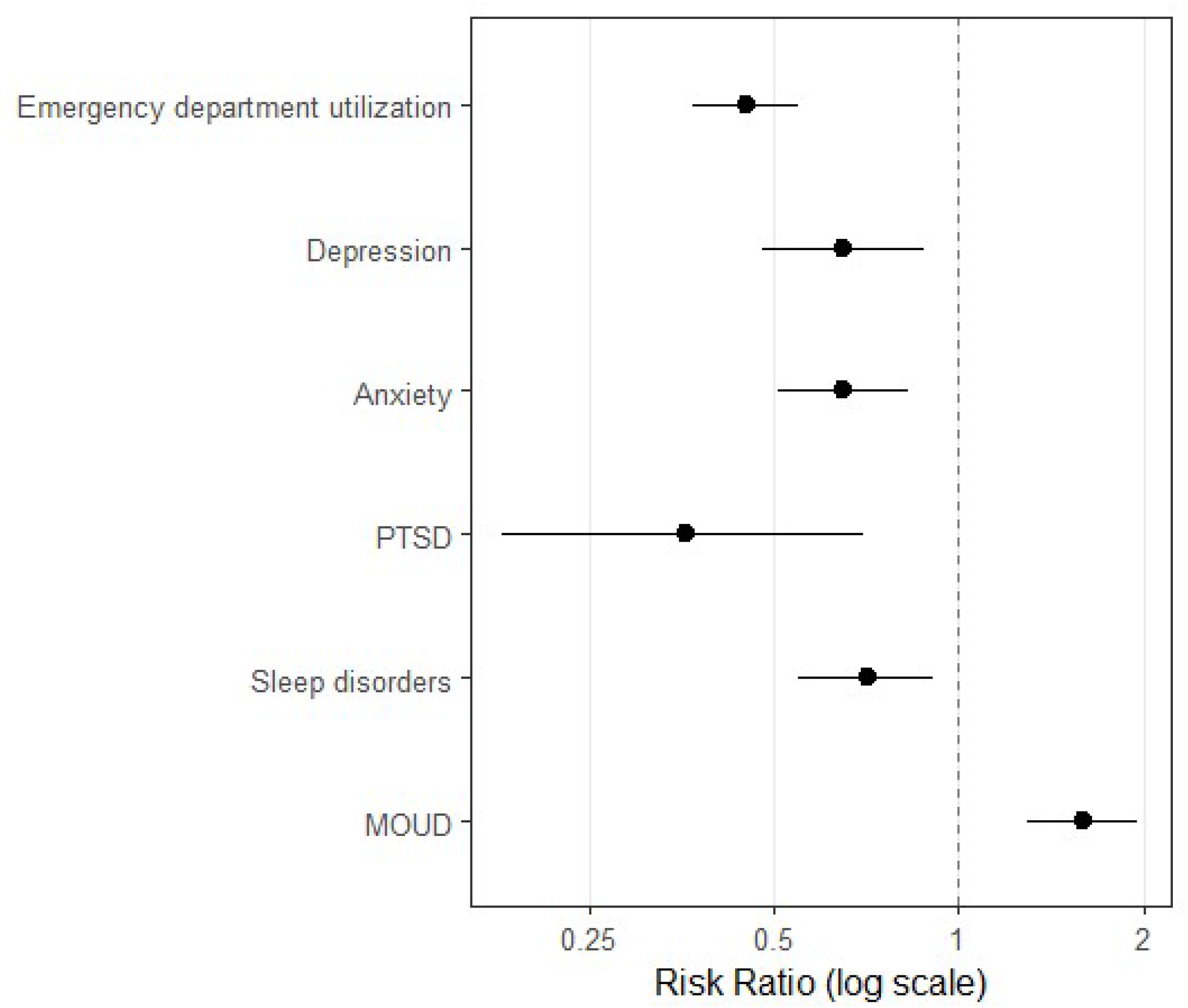
A forest plot of risk ratios (log scale) with 95% CI (confidence intervals) comparing suzetrigine to opioid analgesics for postoperative outcomes. All values less than 1 on the horizontal axis are below the centerline (vertical reference line = 1), indicating that the suzetrigine group has a lower risk for that particular outcome, while all values greater than 1 are above the centerline, indicating that the suzetrigine group had a greater risk for that particular outcome. The width of the error bars represents the 95% CI for each risk ratio.

**Table 1.**
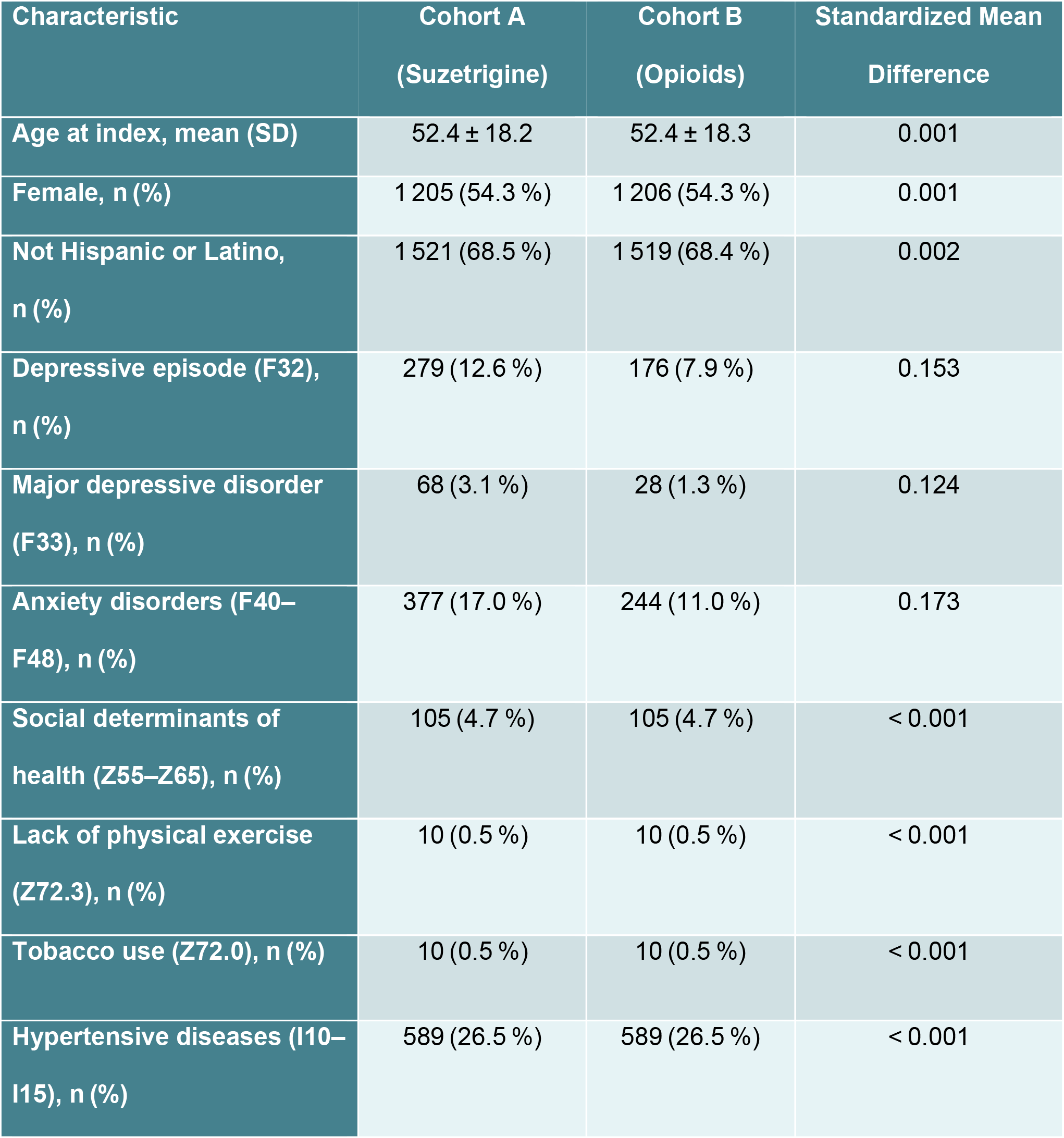
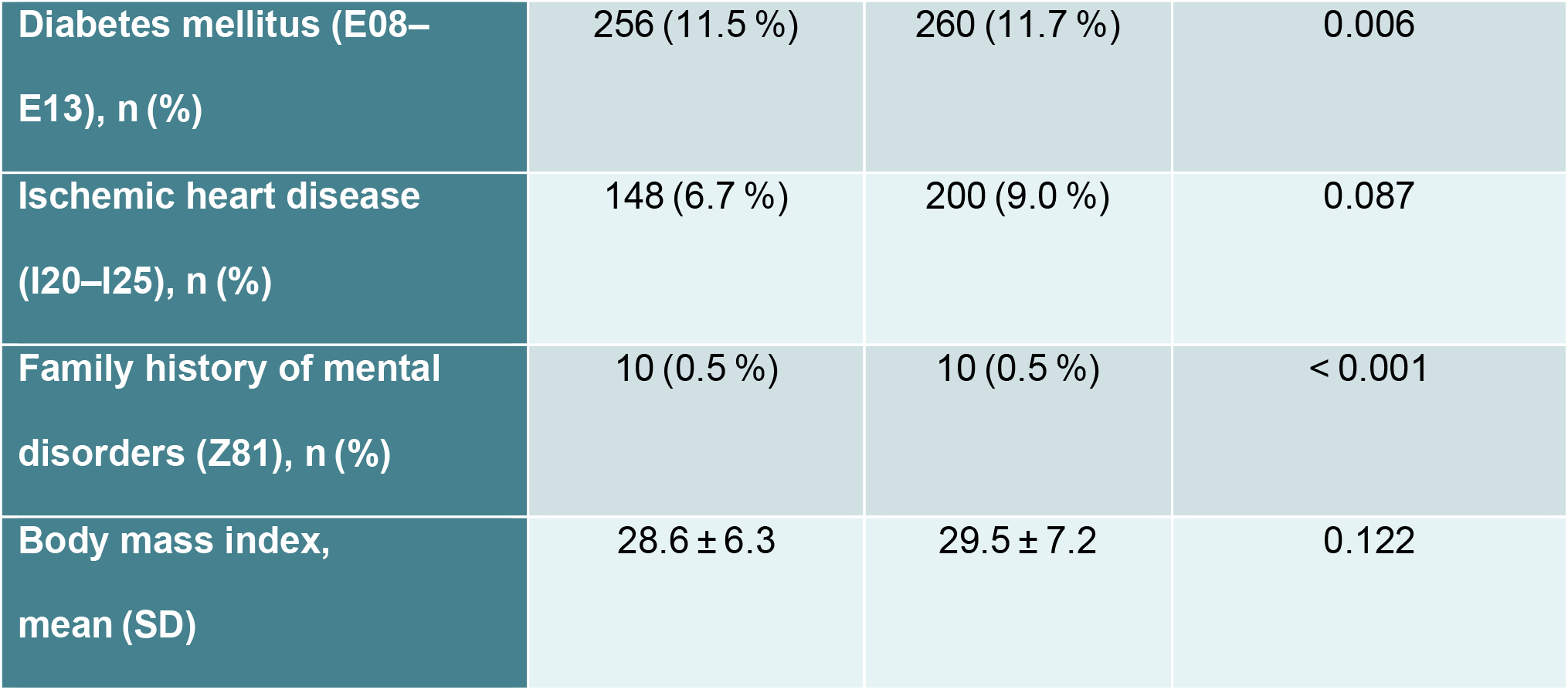
Baseline Characteristics (After Matching)

**Table 2.** Propensity Score Matching Specifications & Balance.

| Item | Value |
| --- | --- |
| Matching ratio | 1:1 (greedy nearest neighbor) |
| Matching algorithm | Greedy nearest neighbor (TriNetX default) |
| Caliper (pooled SD) | 0.1 (TriNetX default) |
| Replacement | Without replacement (TriNetX default) |
| Index date | Date of surgery when exposure criteria met |
| Time window (follow-up) | 1–365 days post-index |
| Propensity score covariates | Age at index; sex; race/ethnicity; social determinants of health (Z55–Z65); lack of physical exercise (Z72.3); tobacco use (Z72.0); hypertensive diseases (I10–I15); diabetes mellitus (E08–E13); ischemic heart disease (I20–I25); family history of mental/behavioral disorders (Z81.x); BMI |
| **Matched sample size** | 2221 per cohort after matching |
| **Balance assessment** | Standardized mean differences < 0.1 for all covariates |

**Table 3.** Clinical Outcomes (Risk Ratios)

| **Outcome** | **Cohort A**<br>**n (%)** | **Cohort B**<br>**n (%)** | **Risk Ratio**<br>**(95 % CI)** | **p-value** |
| --- | --- | --- | --- | --- |
| **Emergency department utilization** | 132 (5.9 %) | 292 (13.1 %) | 0.45 (0.37–0.55) | < 0.001 |
| **Depression** | 68 (3.1 %) | 104 (4.7 %) | 0.65 (0.48–0.88) | 0.005 |
| **Anxiety** | 104 (4.7 %) | 160 (7.2 %) | 0.65 (0.51–0.83) | < 0.001 |
| **Post-traumatic stress disorder (PTSD)** | 11 (0.5 %) | 31 (1.4 %) | 0.36 (0.18–0.70) | 0.002 |
| **Sleep disorders** | 94 (4.2 %) | 133 (6.0 %) | 0.71 (0.55–0.91) | 0.008 |

